# Study protocol: Feasibility and clinical implications of real-time cerebral autoregulation monitoring in major noncardiac surgery with the Medtronic Cotrending algorithm (AUTOREGULATE-NONCARDIAC-COTRENDING)

**DOI:** 10.64898/2026.06.11.26355429

**Authors:** Maximilian Gross, Christian Schindler, Andreas P. Vogt, Urs Pietsch, Miodrag Filipovic, Luzius A. Steiner, Patrick M. Wanner, the Personalising Acute Care Network

## Abstract

**Background:** Perioperative hypotension is associated with postoperative organ injury. However, trials of hypotension avoidance have not found meaningful improvements in postoperative cardiovascular, renal, neurological or functional outcomes. One possible explanation is that organ perfusion depends on patients’ individual autoregulatory ranges. Hence, technology enabling monitoring of the autoregulatory status of vital organs, e.g. the brain, could provide a physiologic basis for personalising of blood pressure targets. However, current established methodologies for monitoring cerebral autoregulation in noncardiac surgery, e.g. the cerebral oximetry index (COx), are limited by performance and usability. The Medtronic Cotrending algorithm has been developed to provide automated, near real-time assessment of cerebral autoregulation. While feasibility was demonstrated in cardiac surgery, its applicability in major noncardiac surgery remains unknown. This study aims to evaluate the technical feasibility and clinical implications of Cotrending-based cerebral autoregulation monitoring in major noncardiac surgery.

**Objectives:** Primary objective: To evaluate the technical feasibility of using the Medtronic Cotrending algorithm to monitor intraoperative cerebral autoregulation in real-time during major noncardiac surgery, drawing comparisons to the COx algorithm. Secondary objectives: to investigate the potential clinical implications of Cotrending-based cerebral autoregulation monitoring.

**Design:** Single-centre, prospective cohort study.

**Setting:** Swiss tertiary care centre

**Patients:** Patients enrolled in AUTOREGULATE-NONCARDIAC who were monitored intraoperatively with the Medtronic INVOS™ 5100 near-infrared spectroscopy (NIRS) system.

**Outcomes:** Technical feasibility outcomes include success rate of determination of the lower limit of cerebral autoregulation, intraoperative uptime, time to first estimate of the lower limit of cerebral autoregulation, sensitivity to external factors and to data artefacts; agreement of Cotrending-derived lower limit of cerebral autoregulation with COx-derived lower limit of cerebral autoregulation.

## BACKGROUND

Intraoperative hypotension is strongly associated with postoperative organ injury.^1^ In the trials to date, various forms of hypotension avoidance have failed to reduce the burden of postoperative organ injury. One compelling explanation is that patients likely differ in their safe blood pressure (BP) ranges, i.e. their autoregulatory ranges.^2,3^ Data from cardiac surgery have shown that BP excursions below the boundaries of cerebral autoregulation are associated with neurological and renal injury, as well as cardiovascular complications, i.e. that the safe BP range for the brain can be used as a surrogate for the safe BP range of other vital organs.^4^

In noncardiac surgery there is a paucity of data on leveraging cerebral autoregulation monitoring, with a few small studies having investigated the use of near-infrared spectroscopy (NIRS)-based cerebral autoregulation monitoring.^5^ The multicentre cohort study AUTOREGULATE-NONCARDIAC is currently investigating the clinical implications and feasibility of cerebral autoregulation-based precision BP monitoring. The current gold standard for NIRS-based cerebral autoregulation monitoring utilises the cerebral oximetry index (COx), an established measure of cerebral autoregulatory function. Despite its strong methodological underpinnings, the COx methodology has two important shortcomings: performance and usability. The generation of reliable estimates of cerebral autoregulatory parameters requires sufficient amounts of data, as well as a certain steady state, a challenge in the dynamic environment of noncardiac surgery evidenced by the high failure rate of COx curve generation described in the literature.^6^ Moreover, interpretation of the derived cerebral autoregulatory curves and parameters requires expertise, making translation to large clinical trials and to clinical practice difficult. Hence, there is a need for novel approaches to cerebral autoregulation monitoring addressing these performance and usability issues. One potentially promising algorithm is the Medtronic Cotrending algorithm, which in cardiac surgery has shown strong agreement with the COx methodology and provided rapid estimates of cerebral autoregulatory state with high uptime and user-friendly visualization of autoregulatory status.^7^

The aim of this sub-study is to investigate the feasibility and impact of the Medtronic Cotrending algorithm. While the Cotrending method has already demonstrated its feasibility in patients undergoing cardiac surgery, further studies are needed to explore its clinical benefits outside of cardiac surgery.^8^ The AUTOREGULATE-NONCARDIAC-COTRENDING substudy aims to address this research gap.

## METHODS

### Study design

AUTOREGULATE-NONCARDIAC-COTRENDING (Clinicaltrials.gov NCT07630129)is an investigator-initiated substudy of the AUTOREGULATE-NONCARDIAC study. The aims and methodology of AUTOREGULATE-NONCARDIAC have been previously described: in summary, it is an investigator-initiated, multicentre prospective cohort study in adult patients at cardiovascular risk undergoing elective major noncardiac surgery, that is investigating the clinical implications and feasibility of cerebral autoregulation monitoring in noncardiac surgery (Clinicaltrials.gov NCT05336864).^9^

### Study setting

AUTOREGULATE-NONCARDIAC-COTRENDING will be conducted at the University Hospital Basel. Enrolment of AUTOREGULATE-NONCARDIAC has concluded (NCT05336864).

### Study population

This substudy includes all patients enrolled in AUTOREGULATE-NONCARDIAC at the University Hospital Basel, where intraoperative cerebral oximetry monitoring was performed using the Medtronic INVOS 5100 system. The eligibility criteria for AUTOREGULATE-NONCARDIAC have been previously described in detail.^9^ In summary, patients 45 years or older, at cardiovascular risk, undergoing major noncardiac surgery with intraoperative invasive blood pressure monitoring, an expected surgical duration of *≥* 90 minutes, and a planned postoperative hospital stay of at least one night are eligible.

### Ethical considerations

Approval for the conduct of this substudy using existing data was granted by the Ethics Committee of Northwestern and Central Switzerland (EKNZ 2025-02371) in December 2025. The current version of the protocol (version 1.0) dates from 15 November 2025. The study is being conducted in accordance with the study protocol, the Declaration of Helsinki, ICH-GCP, and Swiss legal and regulatory requirements for clinical research.

### Objectives

This substudy comprises the following objectives:

#### Objective 1

To evaluate the technical feasibility of real-time cerebral autoregulation monitoring using the Medtronic Cotrending algorithm in patients undergoing major noncardiac surgery, drawing comparisons to other algorithms, including the COx methodology. Feasibility will be assessed looking at the:

- **success rate of cerebral autoregulatory parameter determination**, i.e. the proportion of patients in study cohort in whom cerebral autoregulatory parameters (e.g. the lower limits of cerebral autoregulation [cLLA]) could be determined
- **intraoperative uptime**, i.e. the percentage of intraoperative time during which presumed valid and actionable estimates of cerebral autoregulatory parameters (e.g. cLLA) were delivered
- **time to first cerebral autoregulation parameter estimate**, i.e. the elapsed intraoperative time to first presumed valid estimates of cerebral autoregulatory parameters (e.g. cLLA)
- **sensitivity to external factors**, i.e. the sensitivity of the algorithm to known confounders and determinants of cerebral autoregulatory function, e.g. changes in gas exchange, administration of vasopressors, skin pigmentation.
- **sensitivity to data artefacts**, i.e. the sensitivity of the algorithm to artefacts in data (e.g. in BP or rSO2 signals).

Moreover, the agreement of global estimates of the lower limit of cerebral autoregulation, i.e. estimates of the average lower limit of cerebral autoregulation utilising each patient’s full intraoperatively monitored time, between Medtronic Cotrending and COx will be investigated.

#### Objective 2.1

To determine the extent of between-patient variability in the intraoperative Cotrending-derived boundaries of cerebral autoregulation.

#### Objective 2.2

To determine the extent of within-patient variability in the intraoperative Cotrending-derived boundaries of cerebral autoregulation.

#### Objective 2.3

To determine to what extent the intraoperative Cotrending-derived boundaries of cerebral autoregulation can be predicted using preoperative or preinduction blood pressures.

#### Objective 2.4

To determine to what extent within-patient variability in the intraoperative Cotrending-derived boundaries of cerebral autoregulation can be explained by other intraoperative factors, e.g. CO2 variability.

#### Objective 2.5

To investigate the association of disturbed intraoperative Cotrending-derived cerebral autoregulatory function with the composite outcome of postoperative organ injury (myocardial injury and/or acute kidney injury) on postoperative days 1-3.

#### Objective 2.6

To investigate the association of disturbed intraoperative Cotrending-derived cerebral autoregulatory function with the composite outcome of major cardiovascular, renal and neurological complications up to 1 year following surgery.

### Study methodology

#### Data capture and processing pipeline

The University Hospital Basel is the Data Coordinating Centre. All perioperative data signals relevant to the substudy (e.g. invasive arterial blood pressure [ABP], bilateral cerebral NIRS parameters) are captured at high temporal resolution (i.e. with > 100 Hz sampling rate, enabling reconstruction of physiologic waveforms) using the software ICM+® (Cambridge Enterprise, Cambridge, UK). All data processing takes place offline, following conclusion of all measurements.

#### Cerebral autoregulation calculations

##### Cotrending algorithm

The Cotrending algorithm analyses the dynamic relationship between mean arterial pressure (MAP) and regional cerebral oxygen saturation (rSO2). The method evaluates short data windows of approximately 50 seconds, within which signal gradients of MAP and rSO2 are calculated and compared. According to manufacturer documentation, the autoregulatory state is classified based on the directional relationship between MAP and rSO2 signal trends. The physiological interpretation of these classifications corresponds to pressure-passive versus pressure-independent cerebral perfusion states. Cotrending provides a continuous visual representation of cerebral autoregulatory status over time, permitting estimation of the lower limit of cerebral autoregulation. To simulate real-world conditions, data sets will be streamed to the Cotrending algorithm with calculations only conducted on the data that would have been available up to any given time point.

##### COx algorithm

The implementation of the cerebral oximetry index (COx) algorithm in AUTOREGULATE-NONCARDIAC has been previously described in detail.^9^ In summary, COx is an established and validated measure of cerebral autoregulatory function and corresponds to the correlation of MAP and rSO2 in 5-minute time windows and is updated every 60 seconds.^10^ The resulting 1-minute estimates of COx and MAP are used to generate plots visualising the relationship between autoregulatory function (COx) and MAP. In these plots, the lower and upper limits of cerebral autoregulation correspond to the transition points of COx from < 0.3. to > 0.3.

### Statistical analysis plan

#### Sample

The study uses a convenience sample of all eligible patients within the AUTOREGULATE-NONCARIDAC cohort fulfilling the eligibility criteria for this substudy and with intraoperative data.

#### Datasets and handling of artefacts

The following datasets will be used as specified specifically for each objective:

- Raw dataset: original intraoperative dataset with signals arterial BP (ABP) and rSO2, <u>without</u> any artefact removal
- Cleaned dataset: intraoperative dataset with signals ABP and rSO2 that has undergone manual and additional automatic artefact removal

Manual artefact removal entails human inspection of the ABP and rSO2 time trends and removal of any sections of data deemed to be artefactual or physiologically implausible. Automatic artefact removal involves enforcement of the following hard boundaries: ABP signal 0-250 mmHg, arterial pulse pressure 15-200 mmHg, diastolic BP 15-200 mmHg, rSO2 20-100%.

### Planned statistical analyses

#### Objective 1

The **success rate** of determination of the lower limit of cerebral autoregulation, intraoperative **uptime** and **time to first** estimate of the lower limit of cerebral autoregulation will be summarized using means with standard deviations or medians with interquartile ranges, as appropriate, and proportions with 95% confidence intervals. Sensitivity to external factors and to data artefacts will be assessed using descriptive and regression analyses. These analyses will use the <u>raw</u> dataset. Moreover, to evaluate whether any “look-ahead” has occurred during the Cotrending analyses, sensitivity analyses utilising only certain subsets of the datasets will be conducted (e.g. comparison of cLLA estimate at 15 minutes from whole dataset with cLLA estimate from limited dataset with data from 0-15 minutes).

The **agreement** of estimates of the lower limit of cerebral autoregulation between Medtronic Cotrending and COx will be assessed using scatterplots, Bland–Altman analyses, and mixed-effects models with determination of intraclass correlation coefficients (ICC). For the primary agreement analysis, Cotrending will be run on the <u>raw</u> dataset and COx on the <u>cleaned</u> dataset. A sensitivity analysis will be performed running both Cotrending and COx on the <u>cleaned</u> dataset.

#### Objective 2.1

Global Cotrending-derived estimates of the lower limit of cerebral autoregulation, i.e. estimates of the average lower limit of cerebral autoregulation utilising each patient’s full intraoperatively monitored time, will be characterised using descriptive statistics and visualised using density plots. This analysis will use the <u>raw</u> dataset.

#### Objective 2.2

The extent of within-patient variability in Cotrending-derived lower limit of cerebral autoregulation will be characterised using descriptive statistics. Exploratory analyses will investigate the time trends of Cotrending-derived estimates of the lower limit of cerebral autoregulation. This analysis will use the <u>raw</u> dataset.

#### Objective 2.3

The relationship between preoperative blood pressures and the global Cotrending-derived lower limit of cerebral autoregulation will be assessed visually using scatter plots and analytically using linear regression models involving non-linear terms of pre-operative blood pressure and other baseline characteristics (e.g. age), if indicated. This analysis will use the <u>raw</u> dataset.

#### Objective 2.4

Potential factors associated with within-patient variability in the Cotrending-derived lower limit of cerebral autoregulation will be explored using multivariable linear regression models with robust variance estimates. This analysis will use the <u>raw</u> dataset.

#### Objective 2.5

Multivariable logistic regression will be used to investigate the association of intraoperative area under threshold (AUT) with MAP under the Cotrending-derived lower limit of cerebral autoregulation with the composite outcome of perioperative myocardial injury and/or acute kidney injury on postoperative days 1-3, adjusting for age, preoperative high-sensitivity troponin T, Revised Cardiac Risk Index and anaesthetic duration. This analysis will use the <u>raw</u> dataset.

#### Objective 2.6

Multivariable Cox regression will be used to investigate the association of intraoperative area under threshold (AUT) with MAP under the Cotrending-derived lower limit of cerebral autoregulation with the composite of time to major cardiovascular, renal and neurological complications up to 1 year following surgery, adjusting for age, preoperative high-sensitivity troponin T, Revised Cardiac Risk Index and anaesthetic duration. This analysis will use the <u>raw</u> dataset.

### Missing data

#### Intraoperative physiological data

The handling of data artefacts has been described in the Section *Data and handling of artefacts*. No data imputation will be used for missing intraoperative data.

#### Agreement analyses

Agreement between the Cotrending- and COx-derived lower limits of cerebral autoregulation will be assessed only in patients with valid lower limit of cerebral autoregulation estimates from both methods. Patients with valid Cotrending-based estimates of the lower limit of cerebral autoregulation but non-determinable lower limits of cerebral autoregulation using COx (or vice versa) will be reported separately as part of the feasibility assessment.

#### Outcome data

For analyses relating to the composite outcomes perioperative myocardial injury and/or acute kidney injury on postoperative days 1-3, and time to major cardiovascular, renal and neurological complications up to 1 year following surgery, complete-case analyses will be performed.

### Study status

The parent study AUTOREGULATE-NONCARDIAC has completed recruitment (NCT05336864). Data processing in preparation for this substudy, i.e. relating to the COx algorithm, has begun. The Medtronic Cotrending algorithm has not yet been run on any data from AUTOREGULATE-NONCARDIAC, nor has Medtronic had access to any data from AUTOREGULATE-NONCARDIAC.

## DISCUSSION

### Rationale

Despite the strong association of intraoperative hypotension with postoperative organ injury and other complications, trials of hypotension avoidance have failed to consistently reduce the incidence of postoperative cardiovascular, renal and neurological complications.^1,11–17^ One potential explanation for these findings is that patients differ in their autoregulatory ranges and hence in the BP thresholds below which organ injury begins to occur.^2,18,9^ Cerebral autoregulation monitoring represents a promising approach to individualising intraoperative blood pressure management.^9,19^ However, established methodologies such as the COx approach have high failure rates in noncardiac surgery and require expertise to interpret their findings, complicating their translation to clinical trials and clinical practice.

The Medtronic Cotrending algorithm has been developed to provide an automated, user-friendly, and near real-time assessment of autoregulatory status based on routinely available intraoperative signals. By presenting results in an intuitive visual format not requiring any expertise to interpret, Cotrending has the potential to facilitate clinical adoption of autoregulation-guided haemodynamics. However, before this algorithm may potentially be translated to clinical trials and clinical practice in noncardiac surgery, robust studies investigating the algorithm’s performance, accuracy and the potential prognostic implications of the derived autoregulatory parameters are needed. This substudy will help to fill this research gap and potentially inform future research relating to the Cotrending algorithm.

### Population

Patients undergoing major noncardiac surgery are at high risk of intraoperative hypotension and of perioperative myocardial injury, acute kidney injury, and long-term cardiovascular and neurological complications. Given the strong prognostic implications of intraoperative hypotension, this is a clinically meaningful setting to investigate the potential implications of individualised blood pressure management strategies.

### Limitations

Several important limitations should be considered. First, although during our offline analyses data will be streamed to the Medtronic Cotrending algorithm to simulate real-world conditions and further efforts will be made to ensure that no data “look-ahead” occurs during the analyses, this does not guarantee that our findings will replicate in real-world settings. Second, agreement analyses will be performed against COx-derived autoregulatory parameters. Although the COx algorithm is established and validated, it has important methodological and technical limitations. Specifically, its use in noncardiac surgery is associated with a high failure rate, i.e. in a relevant proportion of patients, credible COx curves and hence credible estimates of the boundaries of cerebral autoregulation may not be determined.^6^ As such, although COx is currently considered the gold standard for NIRS-based monitoring of cerebral autoregulatory function, it is an imperfect comparator. Accordingly, the present study evaluates methodological agreement rather than agreement with a physiological gold standard. Third, determination of autoregulatory limits depends on sufficient spontaneous haemodynamic variability during surgery. In some patients, autoregulatory thresholds may not be identifiable. In addition, certain limitations relate to the Cotrending algorithm itself. The algorithm is proprietary, and its internal processing logic is not publicly available. Consequently, the present study evaluates the algorithm’s output rather than its underlying computational methodology. Finally, while feasibility and methodological agreement are necessary prerequisites for clinical implementation, they do not imply that Cotrending-guided blood pressure management will improve patient outcomes – a research question that can only be addressed in an adequately designed randomised controlled trial.

### Potential impact

Cerebral autoregulation-guided blood pressure management is an appealing, physiologically grounded strategy to individualize perioperative haemodynamic targets. In contrast to fixed population-based thresholds (e.g., MAP *≥* 65 mmHg), autoregulation-based approaches aim to maintain arterial pressure within a patient-specific range that preserves vital organ perfusion. However, despite promising data from other clinical settings, e.g. neurointensive care and cardiac surgery, cerebral autoregulation monitoring has yet to be translated to clinical trials and clinical practice in noncardiac surgery. Two major obstacles faced by current cerebral autoregulation monitoring algorithms such as COx are 1) high failure rates for estimation of the boundaries in real-world noncardiac surgical settings and 2) the need for expertise to interpret the credibility of the calculated cerebral autoregulation data.^6^

The Medtronic Cotrending algorithm has the potential to overcome these barriers by providing automated, continuously updated autoregulatory assessments. If shown to be technically feasible and sufficiently concordant with established methodologies, Cotrending could enable practical implementation of individualized blood pressure management strategies in major noncardiac surgery.

## Data Availability

Not applicable (study protocol paper)

## Abbreviations

ABP: arterial blood pressure
AUT: area under threshold
BP: blood pressure
cLLA: cerebral lower limit of autoregulation
COx: cerebral oximetry index
MAP: mean arterial pressure
NIRS: near-infrared spectroscopy
rSO2: regional cerebral oxygen saturation

## Acknowledgments relating to this article

The authors would like to thank all patients who through their participation have made this study possible.

## Role of Medtronic in this substudy

Medtronic is not providing any compensation in any form for the conduct of this substudy. Medtronic had no role in the study design or data collection. Apart from running the Medtronic Cotrending algorithm on the planned datasets on-site at the University Hospital Basel and providing the AUTOREGULATE-NONCARDIAC investigators with the raw and summarised algorithm output, Medtronic will not be involved in any other aspects of data analysis. Medtronic will not be involved in the interpretation of the data or writing of the manuscript. Any publications presenting results from this substudy will be subject to review by Medtronic to a) check the technical accuracy regarding Medtronic products; b) check whether confidential information has been disclosed and c) check that the source of funding or support for the research project has been fully and adequately disclosed. Medtronic will have the right to request reasonable modifications in relation to (a) to (c) of any manuscript or other materials to be published or presented. The ultimate decision relating to the content and dissemination of any publications resulting from this substudy lies with the principal investigator (PMW).

## CRediT Taxonomy

Conceptualisation: PMW, LAS, MG.

Funding acquisition: PMW, LAS.

Methodology: PMW, LAS, CS, MG.

Project administration: PMW, MG.

Supervision: PMW, LAS.

Writing – original draft: MG, PMW, CS.

Writing – review & editing: MG, PMW, CS, APV, UP, MF, LAS.

## Dissemination

The results of this study will be presented at international scientific conferences and published in peer-reviewed journals, as well as posted in plain language on the Personalising Acute Care Network website (https://www.pac-network.org/autoregulate-noncardiac).

## Study organisation

Principal investigator: PMW

Sponsor: LAS

## Supplemental material

The substudy protocol (v.1.0, 15 November 2025) and further information on AUTOREGULATE-NONCARDIAC can be found on the Personalising Acute Care Network website (https://www.pac-network.org/autoregulate-noncardiac).

